# Hypertrophic Cardiomyopathy – The Impact of Age at Diagnosis of the Proband on Genetic Yield, Clinical Presentation, Outcomes, and Yield of Family Screening

**DOI:** 10.1101/2025.04.10.25325621

**Authors:** Elvira Silajdzija, Christoffer Rasmus Vissing, Emma Basse Christensen, Helen Lamiokor Mills, Thilde Olivia Kock, Lars Juel Andersen, Martin Snoer, Jens Jakob Thune, Emil Daniel Bartels, Anna Axelsson Raja, Alex Hørby Christensen, Henning Bundgaard

**Author notes:** **Corresponding author** Christoffer Rasmus Vissing, Department of Cardiology, The Heart Centre, Copenhagen University Hospital, Blegdamsvej 9, DK-2100 Copenhagen, Denmark.

## Abstract

**Background:** Hypertrophic cardiomyopathy (HCM) is a complex disease with variable clinical presentation and familial impact. Age at diagnosis may influence phenotypic expression, but it is unclear if age also affects clinical outcomes, genetic findings, and yield of family screening.

**Methods:** Retrospective cohort study of families screened for HCM in Eastern Denmark (2006-2023). Probands were analyzed by age at diagnosis both continuously and in quartiles: 18-45, 46-56, 57-65, and >65 years.

**Results:** 612 probands (62% male, median 56 years, median follow-up 9 years) and 919 relatives (45% male, median 42 years) were studied. A higher proband age at diagnosis was associated with more left ventricular (LV) outflow tract obstruction (OR 1.19/10-years), hypertension (OR 1.57/10-years) and atrial fibrillation (OR 1.24/10-years), but less LV hypertrophy (wall thickness ≥30 mm, OR 0.52/10-years), and ventricular arrhythmias (OR 0.81/10-years). Older age at diagnosis was associated with higher all-cause mortality, but similar cardiovascular mortality. Sarcomere variants were less common in the oldest versus youngest quartile of probands (13 vs. 42%, p<0.001). The yield of family screening at baseline was higher in probands diagnosed younger (OR 1.34 per 10-year decrease). Long-term HCM incidence in relatives was not associated with proband age at diagnosis, and overall yield of family screening was comparable across all proband ages at diagnosis.

**Conclusions:** The probands age at HCM diagnosis influenced clinical and genetic findings but the cardiovascular mortality and the yield of family screening were similar across all ages at diagnosis, supporting the presently recommended follow-up and family screening irrespective of the probands age at diagnosis.

**Clinical perspectives:** *What is new?:*

- The study demonstrates that the age at diagnosis in HCM probands is significantly associated with distinct clinical and genetic profiles.
- It provides novel insights into family screening yield, revealing that despite differences in baseline genetic yield and clinical presentation across age groups, the overall prevalence and incidence of HCM in relatives remain similar regardless of the proband’s age at diagnosis.

*What are the clinical implications?:*

- These findings support the recommendation for consistent family screening in HCM, regardless of the age at which the proband is diagnosed, as the risk for developing HCM appears comparable across all age groups.
- The identification of age-related differences in clinical phenotype and genetic findings could inform tailored risk stratification and management strategies, potentially enhancing personalized care for HCM patients.

## Introduction

Hypertrophic cardiomyopathy (HCM) is the most common hereditary cardiac disease with a prevalence of 1:500-1:200 and can present at any age. ^1, 2^ Sudden cardiac death (SCD) and severe left ventricular hypertrophy tend to present in younger patients, whereas older patients often have milder phenotypes. ^3–6^ Thus, early onset of HCM is often associated with severe clinical adverse outcomes and familial transmission patterns. ^7^ However, the clinical presentation and the genetics in patients diagnosed at older ages remain underexplored.

The number of older individuals being diagnosed with HCM is increasing and further characterization of presentation and yield of family screening in patients with HCM diagnosed at older age is of interest. ^6, 8, 9^ Hypertension, valvular dysfunction, chronic obstructive pulmonary disease, diabetes etc. are more common in older patients and these comorbidities may influence the cardiac phenotype in patients with HCM. ^10^

Previous reports have suggested that HCM patients diagnosed at older age have a lower prevalence of family history of HCM compared to HCM patients diagnosed younger. ^11–13^ It remains unclear whether screening and continued surveillance for HCM is as warranted in relatives of probands diagnosed at older age as in those from families with probands diagnosed at younger age.

In this study, we characterized the presentation and disease course of HCM in probands according to age at diagnosis. In addition, we assessed the associated diagnostic yield of family screening according to proband’s age at diagnosis.

## Methods

### Study design and population

This was a retrospective, longitudinal, observational cohort study comparing adult probands diagnosed at different ages. The study population was divided into quartiles, resulting in the following age groups: 18-45 years, 46-56 years, 57-65 years, and >65 years. All probands were evaluated and followed between 2006 and 2023 at a regional assembly of clinics for inherited cardiac diseases covering Eastern Denmark (population ∼2.8 million). According to international ^12, 14, 15^ and Danish ^16, 17^ guidelines, the diagnostic criteria for HCM in probands and relatives were maximal left ventricular wall thickness (MWT) ≥15 mm and ≥13 mm, respectively, in the absence of other explanatory cardiac or systemic diseases.

Probands with confirmed or suspected HCM were referred by general practitioners, private cardiologists, or local cardiology clinics. At the initial consultation of the HCM proband, a three-generation family pedigree was drawn to identify relatives at risk and cascade screening was commenced.

For this study all patients with HCM were characterized phenotypically and we collected data on demographics, symptoms, medical and family history, genetics, electrocardiogram, arrhythmia monitoring, transthoracic echocardiography, and adverse outcomes from electronic medical records and the national pedigree database Progeny (version 9, Progeny Genetics LLC, Aliso Viejo, California).

### Outcomes

Studied outcomes included atrial fibrillation/flutter, ventricular arrhythmias (sustained ventricular tachycardia, ventricular fibrillation, aborted sudden cardiac death defined as resuscitated cardiac arrest, sudden cardiac death or appropriate anti-tachycardia pacing and/or shock therapy), left ventricular systolic dysfunction (left ventricular ejection fraction <50%), septal reduction therapy, stroke, heart transplantation, all-cause mortality, and cardiovascular death.

Data was collected from the first screening visit and from the most recent visit in the study period.

### Genetic screening

All probands with HCM were offered genetic screening, and all relatives were offered genetic testing if they derived from a family with a likely pathogenic (LP) or pathogenic (P) genetic variant according to the American College of Medical Genetics and Genomics (ACMG) criteria. ^18^

The list of HCM-associated genes sequenced throughout the study period is provided in Supplementary Methods. Initially, Sanger sequencing was used for genetic screening, but later next generation sequencing with larger panels was introduced, resulting in an evolution of the number of screened genes. At the end of the study period, all genetic results were re-evaluated and re-classified according to the ACMG criteria. ^18^

Probands without identifiable sarcomere genetic variants, or carrying variants of unknown significance (VUS), were categorized as gene elusive. Individuals with LP/P variants in *GLA*, *PTPN11* or in genes predominantly associated with other phenotypes (e.g., *RBM20, DES* and *SCN5A*) and with phenocopies of HCM, e.g., amyloidosis, were excluded. Relatives of probands not genetically tested were also excluded from the analyses.

### Statistical analyses

Categorical variables are presented as counts and percentages, and continuous variables as mean ± standard deviation (SD) or median with interquartile range (IQR), according to data distribution. Groups were compared using Chi-Squared Test, Fisher’s exact test, or Kruskal-Wallis rank sum test, as appropriate. The Kaplan Meier method was used to estimate the cumulative incidence of adverse outcomes in probands stratified according to age at diagnosis and compared using a log-rank test. Proband age at HCM diagnosis was both investigated as a continuous variable and grouped into four quantiles (18-45, 46-56, 57-65 and >65 years). We investigated the association between age at diagnosis and adverse outcomes during follow-up in HCM probands using a Cox proportional hazards model approach. In Cox proportional hazards modeling, the follow-up period from the first screening visit was used as the timescale. Descriptive analyses and age-specific incidence rates in relatives were reported according to the same grouping of age as in probands, i.e., 18-45 years, 46-56 years, 57-65 years, and >65 years.

The default reported confidence interval (CI) was 95% CI, and level of statistical significance was set at p <0.05. All statistical analyses were performed in R, version 4.3.1 (R Foundation for statistical computing, Vienna, Austria), including use of the packages tidyverse, survminer, survival, and gtsummary. The data underlying this paper are not publicly available because of national personal data protection regulations.

### Ethics

The study was approved by the Danish Agency for Patient Safety, and the Danish Data Protection Agency with case number 3-3013-2380/1 and was conducted according to the Declaration of Helsinki.

## Results

### Study population

We assessed probands from 660 families for potential inclusion in the study. After excluding phenocopies and probands <18 years of age at diagnosis, 612 adult probands were included in the study. The age and sex distributions are seen in Table 1.

**Table 1:**
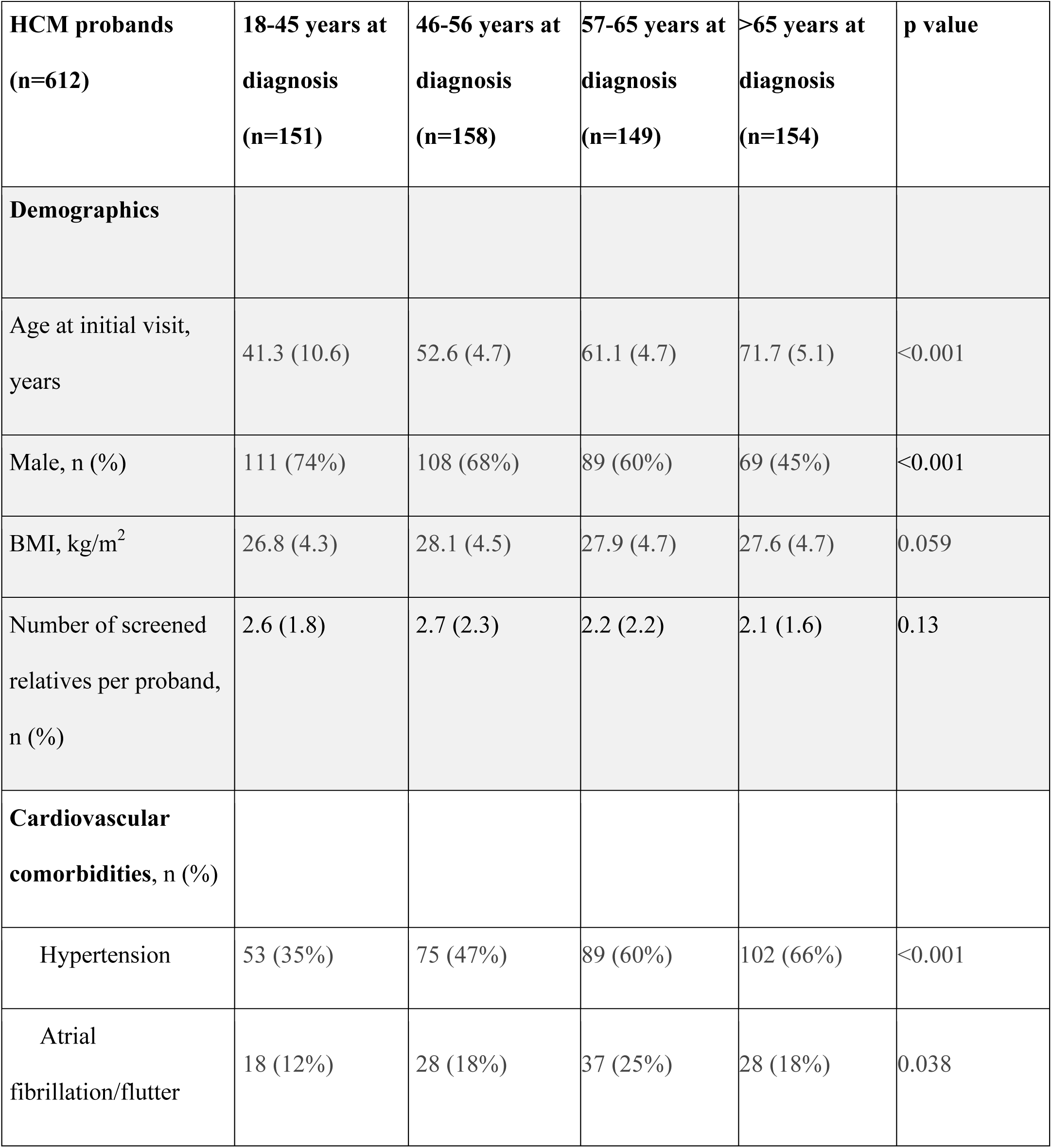
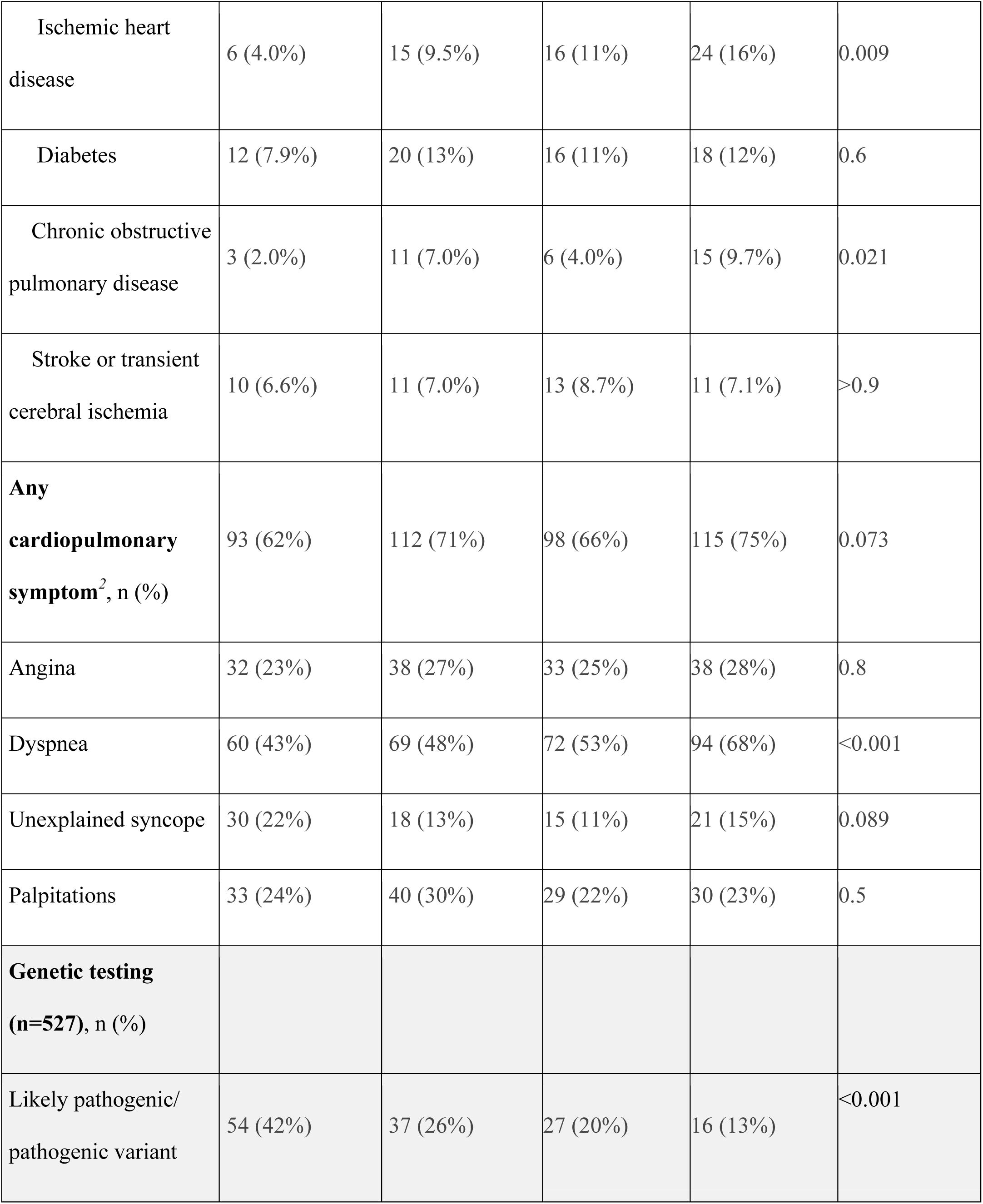
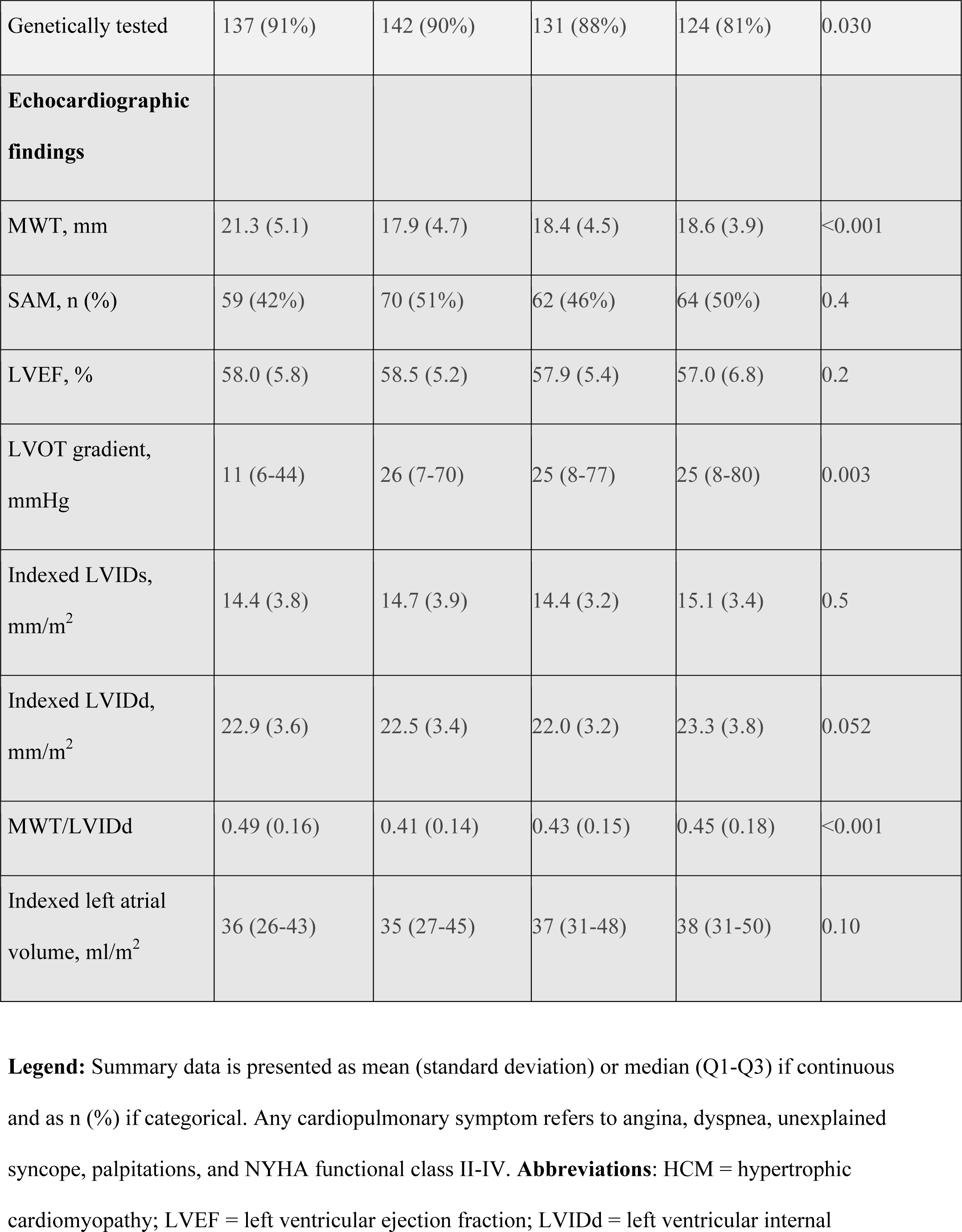
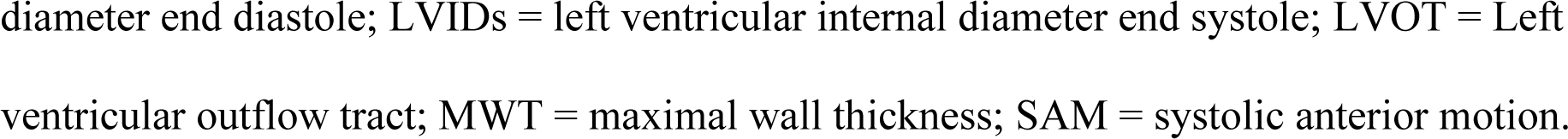
Clinical characteristics in HCM probands at baseline (n=612) stratified according to age at diagnosis. ^1^

### Clinical characteristics of probands according to age at diagnosis

Median age at HCM diagnosis was 56 (IQR 46-65) years, with a median follow-up time of 8.7 (IQR: 5.1-12.6) years. Clinical characteristics according to age at diagnosis in probands, corresponding to the quartiles at diagnosis are provided in Table 1. Females were diagnosed with HCM later in life than males (mean 59 vs. 53 years, p<0.001). Probands diagnosed older had a significantly higher prevalence of hypertension, ischemic heart disease, and chronic obstructive pulmonary disease compared to probands diagnosed at younger ages (Figure 1, Table 1). At baseline, probands diagnosed >65 years experienced more dyspnea, but otherwise the symptom burden was comparable across all ages at diagnosis (Table 1). Differences in baseline echocardiographic parameters included an association between younger age at diagnosis and severe LV hypertrophy (defined as LV wall thickness of ≥30 mm; OR 0.52 per 10 years, Figure 1), with significantly less left ventricular hypertrophy (LVH) in the oldest (>65 years) compared to the youngest (18-45 years) ages at diagnosis (21.3±5.1 mm vs. 18.6±3.9 mm, p=0.002; Table 1). In contrast, older age at diagnosis was associated with a higher occurrence of obstructive physiology of the left ventricular outflow tract at baseline (Figure 1), with higher max left ventricular outflow tract gradients in the oldest compared to the youngest ages of diagnosis (25 (IQR: 8-80) vs. 11 (IQR: 6-44) mmHg, p=0.003; Table 1).

**Figure 1:**
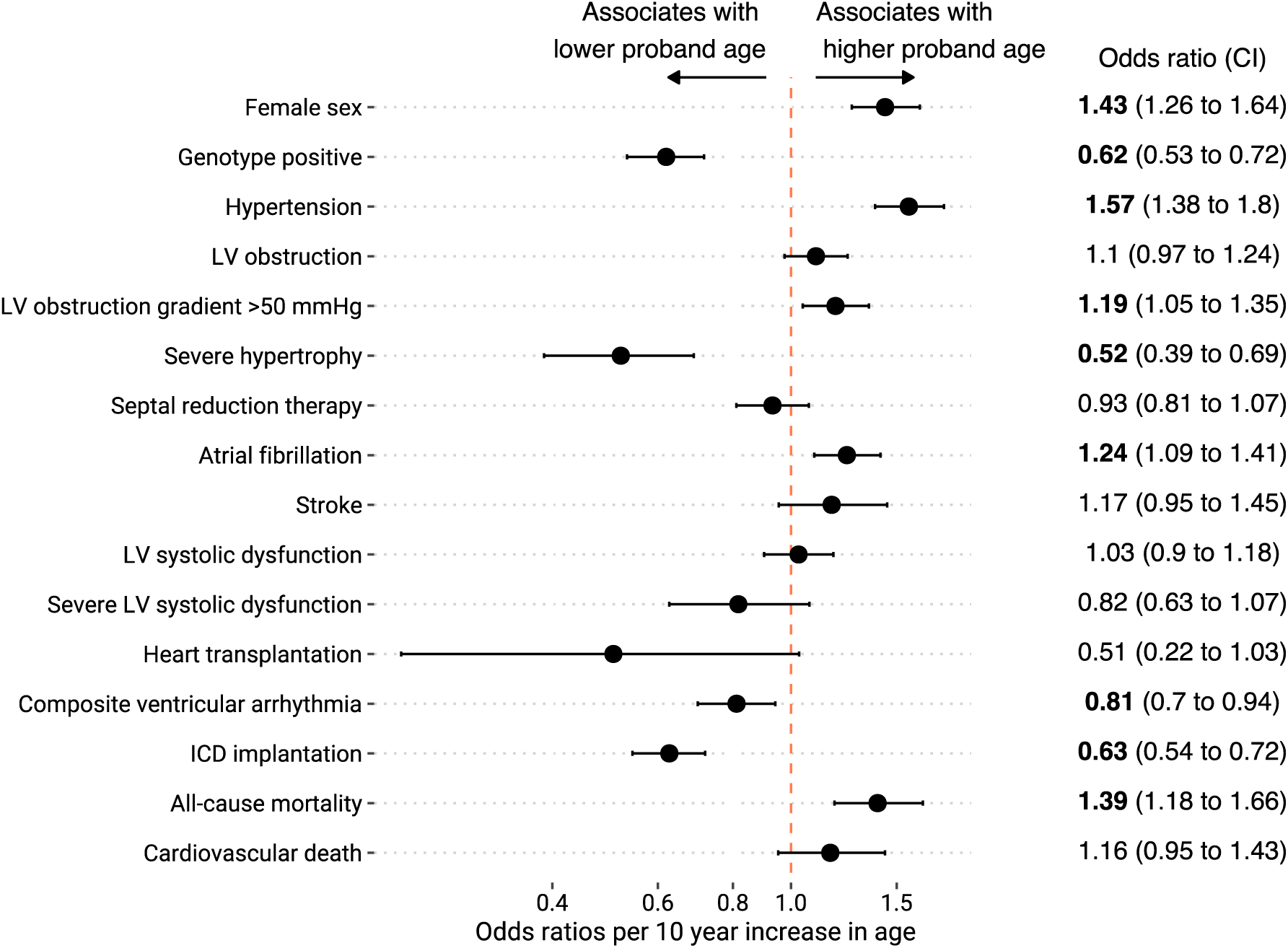
Adverse outcomes in hypertrophic cardiomyopathy probands sub-grouped by age at diagnosis. Adverse outcomes according to proband age at diagnosis represented as odds ratios per 10 years increase/decrease in proband age at diagnosis. Adverse outcomes are as stated in the diagram. Dots denote odds ratio and vertical breaks the confidence interval.

### Genetic findings

Genetic screening had been performed in 89% of probands. Supplementary Table 1 provides a full list of all genetic variants and their frequencies in our cohort. A significant positive association between being diagnosed with HCM at a younger age and carrying a LP/P genetic variant was observed both in regression analysis and according to the four investigated groups of ages at diagnosis (Table 1, Figure 1, and Supplementary Table 2). In linear modelling, a 10-year increase in age at HCM diagnosis was associated with an odds ratio of 0.62 (CI 0.53-0.72) for carrying a LP/P variant, with 13% of probands diagnosed >65 years carrying a LP/P genetic variant compared to 43% of probands diagnosed 18-45 years (p<0.001). LP/P variants in *MYBPC3* were the most common genetic finding across all age groups (Supplementary Table 2).

### Clinical findings and events in probands according to age at diagnosis

Older age at diagnosis of HCM was associated with a higher prevalence of severe LV outflow obstruction (gradient >50 mmHg, OR 1.19 per 10 years increase), atrial fibrillation (OR 1.24 per 10 years increase), and all-cause mortality (OR 1.39 per 10 years increase) at end of follow-up. In contrast, older age at diagnosis was associated with a lower prevalence of the composite ventricular arrhythmia outcome (OR 0.81 per 10 years increase) and implantable cardioverter defibrillator (ICD) implantation (OR 0.63 per 10 years increase). While older age at diagnosis was associated with a higher all-cause mortality, no significant difference according to age at diagnosis was observed for cardiovascular death (Figure 1). Counts and percentages of clinical events are given according to age of diagnosis in probands in Table 2.

**Table 2:**
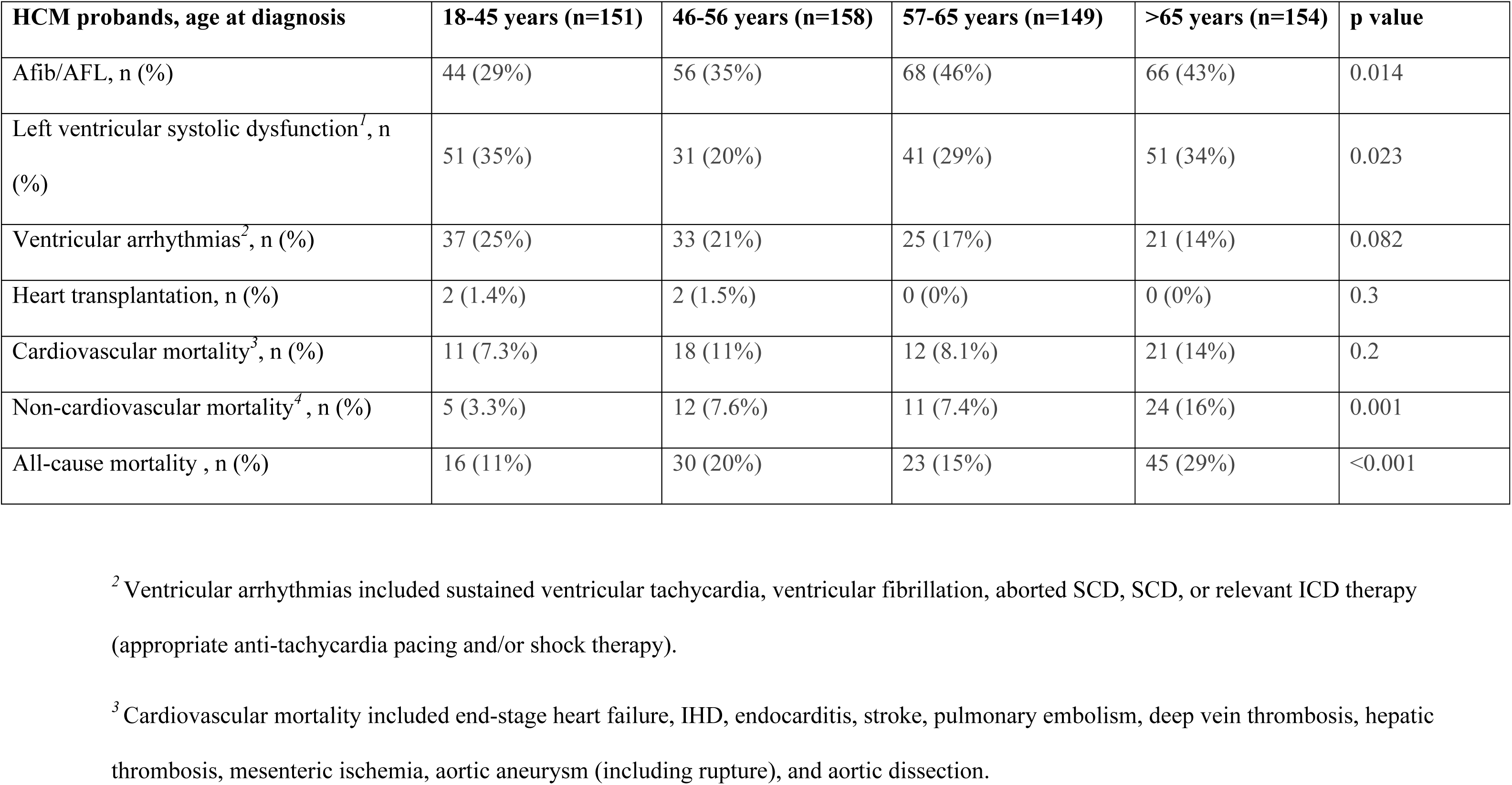
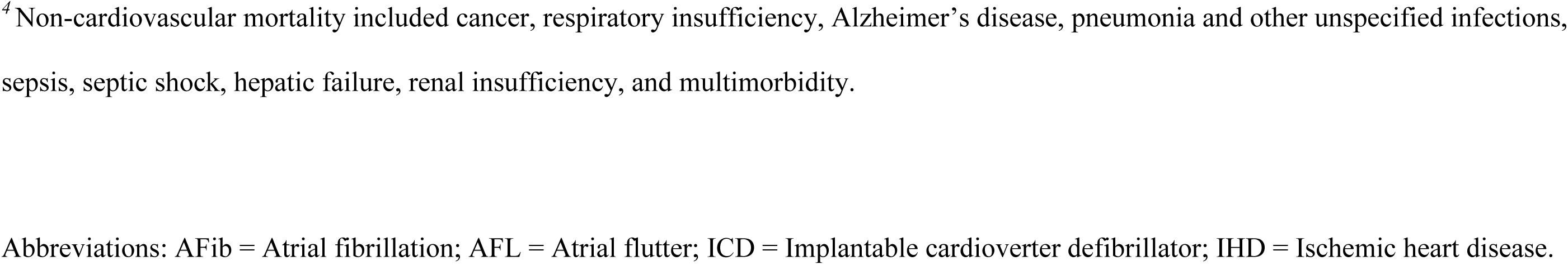
Adverse outcomes at end of follow-up in HCM probands stratified according to their age groups at diagnosis (n=612)

| **HCM probands, age at diagnosis** | **18-45 years (n=151)** | **46-56 years (n=158)** | **57-65 years (n=149)** | **>65 years (n=154)** | **p value** |
| --- | --- | --- | --- | --- | --- |
| Afib/AFL, n (%) | 44 (29%) | 56 (35%) | 68 (46%) | 66 (43%) | 0.014 |
| Left ventricular systolic dysfunction<sup>1</sup>, n (%) | 51 (35%) | 31 (20%) | 41 (29%) | 51 (34%) | 0.023 |
| Ventricular arrhythmias<sup>2</sup>, n (%) | 37 (25%) | 33 (21%) | 25 (17%) | 21 (14%) | 0.082 |
| Heart transplantation, n (%) | 2 (1.4%) | 2 (1.5%) | 0 (0%) | 0 (0%) | 0.3 |
| Cardiovascular mortality<sup>3</sup>, n (%) | 11 (7.3%) | 18 (11%) | 12 (8.1%) | 21 (14%) | 0.2 |
| Non-cardiovascular mortality<sup>4</sup>, n (%) | 5 (3.3%) | 12 (7.6%) | 11 (7.4%) | 24 (16%) | 0.001 |
| All-cause mortality, n (%) | 16 (11%) | 30 (20%) | 23 (15%) | 45 (29%) | <0.001 |

<sup>2</sup> Ventricular arrhythmias included sustained ventricular tachycardia, ventricular fibrillation, aborted SCD, SCD, or relevant ICD therapy (appropriate anti-tachycardia pacing and/or shock therapy).
<sup>3</sup> Cardiovascular mortality included end-stage heart failure, IHD, endocarditis, stroke, pulmonary embolism, deep vein thrombosis, hepatic thrombosis, mesenteric ischemia, aortic aneurysm (including rupture), and aortic dissection.
<sup>4</sup> Non-cardiovascular mortality included cancer, respiratory insufficiency, Alzheimer's disease, pneumonia and other unspecified infections, sepsis, septic shock, hepatic failure, renal insufficiency, and multimorbidity.
Abbreviations: AFib = Atrial fibrillation; AFL = Atrial flutter; ICD = Implantable cardioverter defibrillator; IHD = Ischemic heart disease.

### Yield of family screening

Probands without any screened adult relatives (n=166) or from families not genetically tested (n=29) were excluded from the analyses of family screening. Thus, a total of 919 relatives (median age 42 [IQR 29-55] years, median follow-up 6.4 [IQR: 3.4-10.1] years, 55% female) were included in the analyses. No significant difference was found in number of relatives examined per proband according to proband’s age at diagnosis (Table 3). Relatives of probands diagnosed >65 years of age were significantly older at baseline screening compared to relatives of probands diagnosed at younger ages (Table 3).

**Table 3:** Clinical characteristics at baseline among relatives of probands diagnosed at different ages (n=919)

| **Relatives**<br>**(n=919)** | **Relatives of**<br>**probands 18-**<br>**45 years**<br>**(n=219)** | **Relatives of**<br>**probands 46-**<br>**56 years**<br>**(n=262)** | **Relatives of**<br>**probands 57-**<br>**65 years**<br>**(n=220)** | **Relatives of**<br>**probands >65**<br>**years (n=218)** | **p value** |
| --- | --- | --- | --- | --- | --- |
| **Demographic**<br>**parameters** |  |  |  |  |  |
| Age at initial visit,<br>years | 36.4 (21.5-55.1) | 39.1 (26.6-53.8) | 39.5 (28.5-50.3) | 48.4 (41.0-57.4) | <0.001 |
| Follow-up time,<br>years | 5.0 (1.2, 9.0) | 5.0 (1.8, 9.7) | 5.5 (2.2, 9.4) | 4.5 (1.6, 7.5) | 0.059 |
| Male | 105 (48) | 116 (44) | 99 (45) | 90 (41) | 0.58 |
| BMI, kg/m<sup>2</sup> | 23.2 (19.5-28.0) | 24.2 (21.8-27.8) | 23.9 (21.3-27.7) | 24.3 (21.5-27.1) | 0.19 |
| HCM diagnosed | 40 (18.3%) | 43 (16.4%) | 13 (5.9%) | 26 (11.9%) | <0.001 |
| **TTE findings** |  |  |  |  |  |
| MWT, mm | 10.5 (4.4) | 10.1 (3.7) | 9.4 (2.3) | 9.7 (2.5) | 0.63 |
| SAM, n (%) | 12 (6.4%) | 15 (6.5%) | 2 (1.0%) | 7 (3.6%) | 0.020 |
| LVEF, % | 59.4 (3.9) | 59.6 (2.3) | 59.6 (2.6) | 59.6 (2.7) | 0.90 |
| LVOT gradient,<br>rest, mmHg | 5.0 (4.0-7.0) | 5.0 (4.0-7.0) | 5.0 (4.0-7.0) | 6.0 (4.0-7.0) | 0.52 |
| LVOT gradient,<br>Valsalva, mmHg | 6.0 (4.0-7.0) | 5.0 (4.0-7.0) | 5.0 (4.0-7.0) | 6.0 (4.0-7.0) | 0.40 |
| MWT/LVIDd | 0.23 (0.11) | 0.22 (0.09) | 0.21 (0.06) | 0.21 (0.07) | 0.81 |
| Indexed left atrial<br>volume, ml/m<sup>2</sup> | 24.4 (19.7-30.6) | 24.5 (19.4-29.7) | 22.7 (18.1-28.3) | 25.7 (20.0-32.5) | 0.086 |

Definitions as in Table 1.
Values in the table are given as counts (percentages) for categorical variables and as mean (standard deviation) or median (first quartile-third quartile) for continuous variables, depending on data distribution.
Abbreviations: HCM = hypertrophic cardiomyopathy; LV = left ventricular; LVEF = left ventricular ejection fraction; LVIDd = left ventricular internal diameter end diastole; LVOT = Left ventricular outflow tract; MWT = maximal wall thickness; SAM = systolic anterior motion; TTE = transthoracic echocardiogram

### Risk of HCM at baseline in relatives according to the proband age at diagnosis

In linear regression analysis, we found a significant association between proband age at HCM diagnosis and diagnostic yield of baseline screening, with an odds ratio of 1.34 (CI 1.16-1.64, p<0.001) per 10 year decrease in proband age at HCM diagnosis, adjusted for the relatives’ age at screening and sex (OR 1.21 [CI 1.06-1.38], p=0.003 in unadjusted analysis) (Table 3). The crude prevalence of HCM in relatives at baseline screening was not significantly different in subgroup analyses of families from probands aged 18-45 vs. >65 years at HCM diagnosis (18% vs 12%, OR 1.65 [CI 0.94-2.94], p=0.08, Figure 2). However, the prevalence of HCM at first screening visit, adjusted for the relative’s age, was markedly higher in relatives from younger probands with a baseline age of 46-65 years in relatives (Figure 2). No significant differences in echocardiographic findings were observed between relatives of probands diagnosed at different ages (Table 3). Similarly, electrocardiographic findings (hypertrophy, 2^nd^/3^rd^-degree AV-block, abnormal repolarization, presence of bundle branch block) across all relatives of probands diagnosed at different ages were similar.

**Figure 2:**
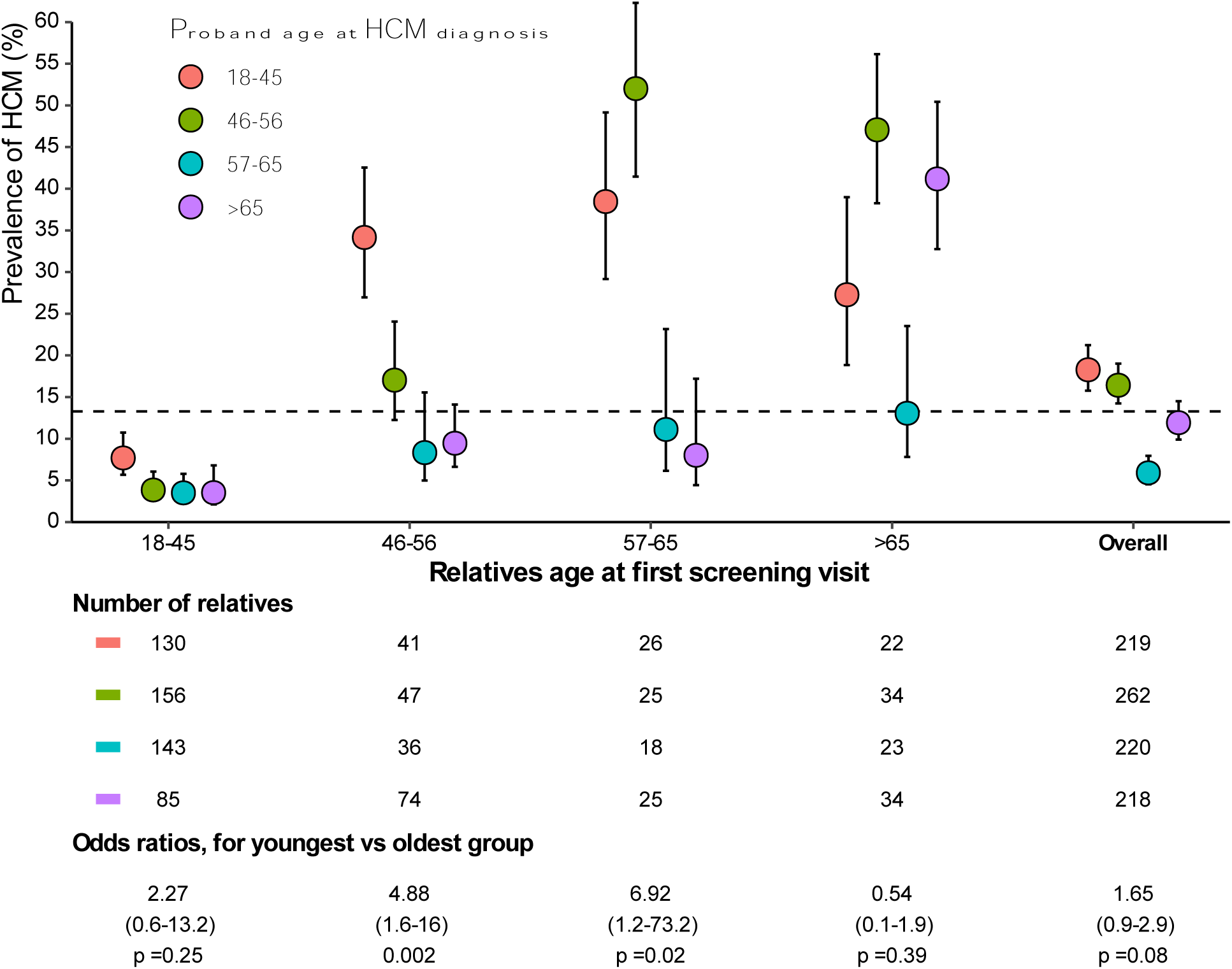
Yield of baseline screening in relatives according to age of relative at time of baseline screening, stratified for age of proband at hypertrophic cardiomyopathy diagnosis. Yield of baseline screening of relatives according to the proband age at diagnosis of hypertrophic cardiomyopathy (HCM) (represented by colors) and age of relative at first screening (x-axis). The number of relatives screened in each group is provided under the plot and the odds ratio comparing the prevalence of HCM at first screening in relatives of probands diagnosed youngest and oldest is also provided.

### Findings in relatives during follow-up according to age of proband

In 797 relatives without HCM at baseline, 37 were diagnosed with HCM during 5,318 patient-years of follow-up. No difference in the incidence of HCM was observed among relatives to probands diagnosed at different ages (log-rank p=0.28), or when using proband age as a continuous variable in Cox proportional hazards modeling (HR 0.98 per 10 years, p=0.82). The overall proportion of relatives diagnosed with HCM at end of follow-up in relatives of probands diagnosed at 18-45, 46-56, 57-65, and >65 years of age were 22% (n=47), 22% (n=58), 9% (n=20), and 15% (n=33), respectively (relatives of probands diagnosed 18-45 vs. >65 years, OR 1.53 [CI 0.91-2.59], p=0.11). During follow-up, an association between proband age at diagnosis and the incidence of ICD implantation (HR 0.56 per 10 years, p <0.001) and developing atrial fibrillation (HR 1.42 per 10 years, p=0.046) in relatives was observed with no significant associations between proband age at diagnosis and development of HCM or other adverse outcomes in relatives (Figure 3).

**Figure 3:**
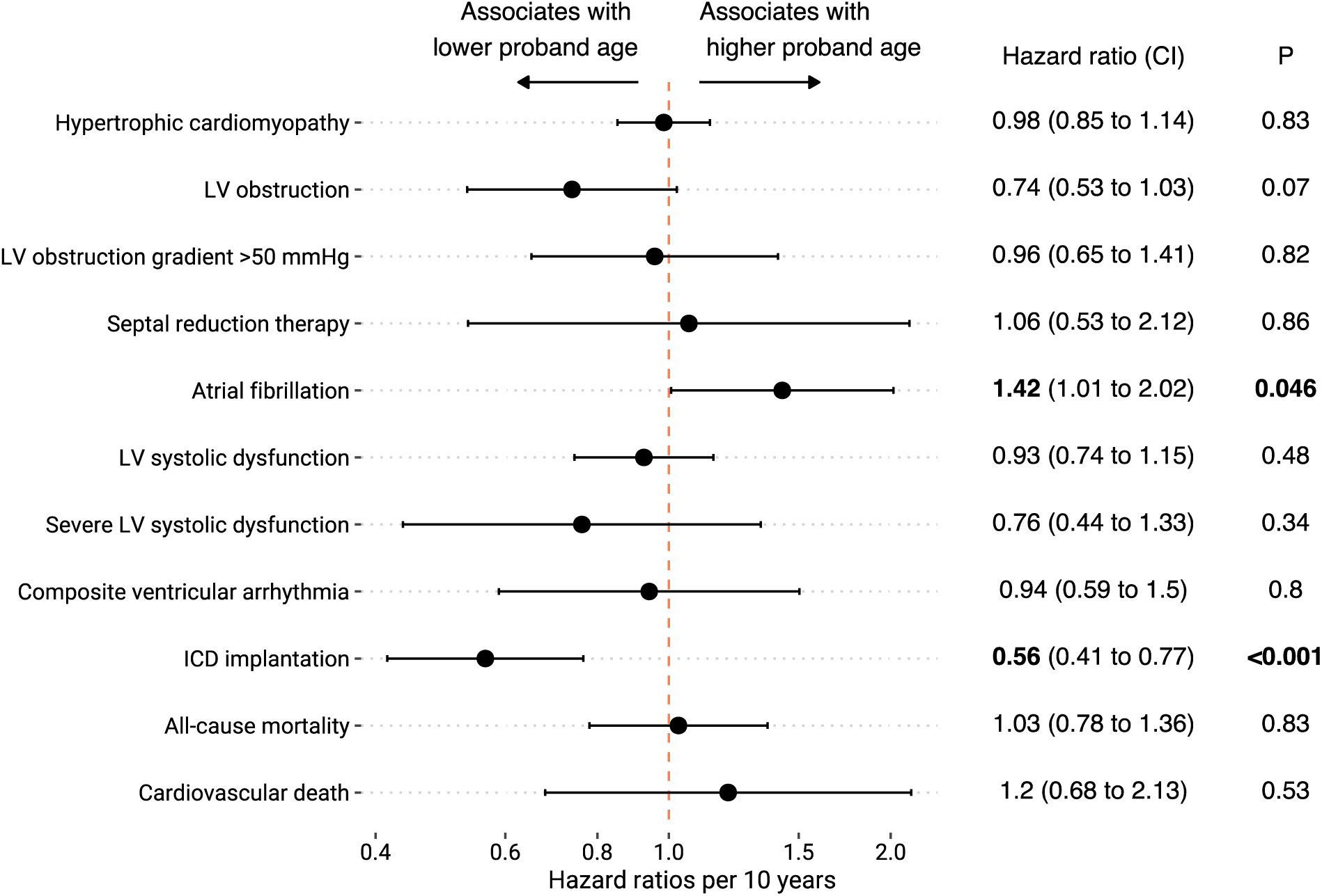
Adverse outcomes during follow-up in relatives according to proband’s age at diagnosis. Forest plot presenting the associations between age at time diagnosis of HCM in the family proband to the incidence of adverse outcomes during follow-up in screened relatives. Hazard ratios from univariate analysis are given per 10-year increase in proband age at diagnosis.

### Findings according to genetic status

The diagnostic yield at baseline screening of relatives to probands with HCM was higher in relatives from genotype positive families compared to relatives to gene elusive probands (18% vs. 11%, OR 1.81 [CI 1.21-2.70], p=0.002; Figure 4A). Further stratification for proband age at diagnosis, identified a significantly higher prevalence of HCM at first screening of relatives to genotype-positive compared to gene elusive probands diagnosed at the youngest age, while no statistically significant differences were observed for genotype-positive or gene elusive probands diagnosed older (Figure 4A). A significant difference in incidence of HCM during follow-up according to genetic findings was observed for probands diagnosed the second-youngest (age 46-56 years) with an incidence ratio of 2.78 (CI 1.33-5.11, p=0.008), while no significant differences were observed for probands diagnosed at other ages or overall (Figure 4B).

**Figure 4:**
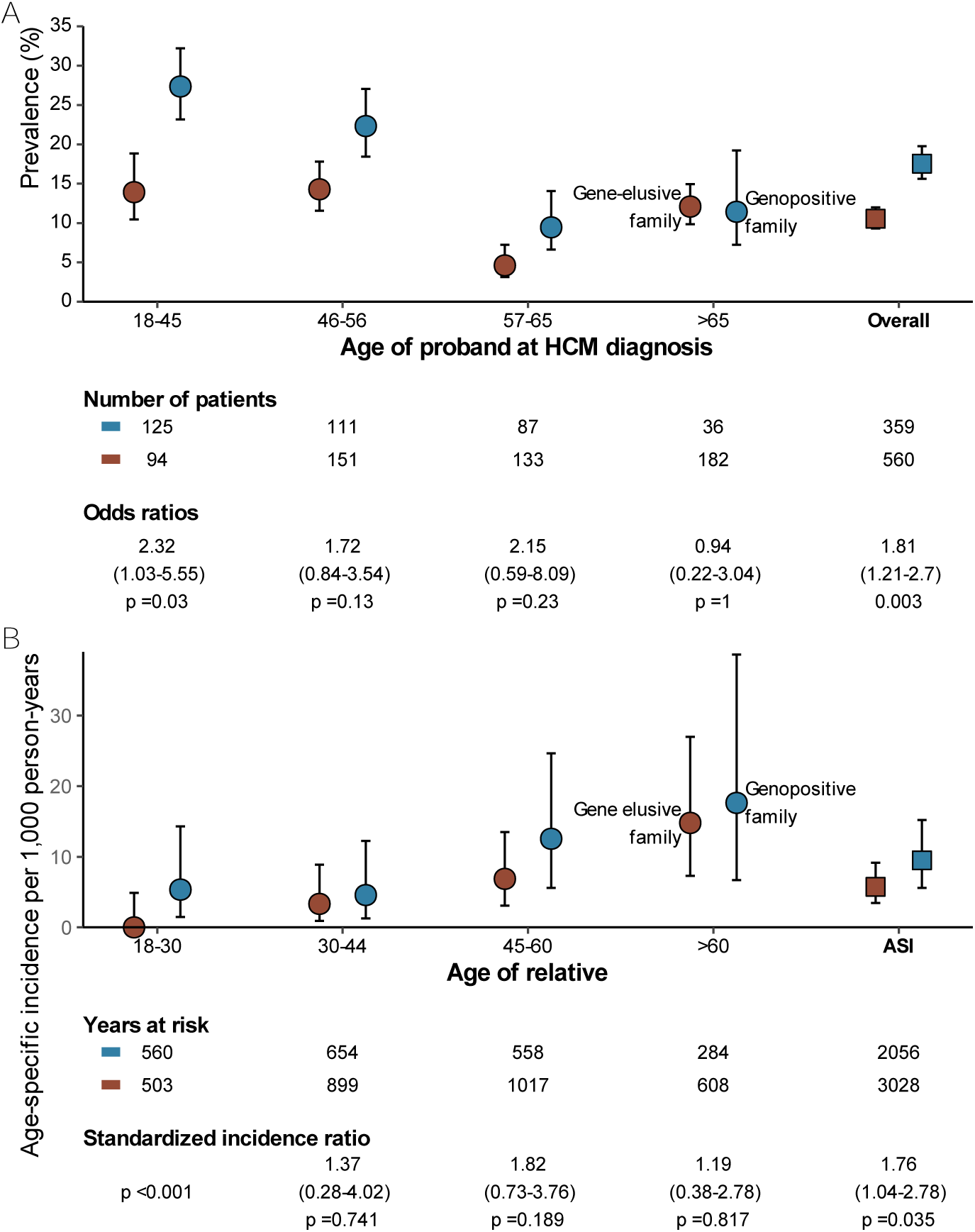
HCM at baseline and during follow-up in relatives of genotype positive and gene elusive probands according to proband’s age at diagnosis. Yield of screening in genotype positive (blue) and gene-elusive (red) families according to the age of the family proband at time of hypertrophic cardiomyopathy (HCM) diagnosis (x-axis). **A**) Prevalence of HCM at first screening visit in relatives. **B**) Incidence of HCM during follow-up in relatives without HCM at baseline screening. The number of patients in each group (A) or total years at risk (B) and statistical comparisons between the groups are given at the bottom of each panel.

## Discussion

In this study, we identified significant associations between proband age at HCM diagnosis and distinct genetic, phenotypic, and clinical outcomes. Older age at HCM diagnosis was associated with a lower yield of genetic testing, lower maximal LV wall thickness, and a lower family screening yield, particularly among younger relatives. In relatives the prevalence of HCM at baseline screening was similar among probands diagnosed at different ages (18% vs. 12% for youngest vs. oldest), and no differences were observed in incidence of HCM during follow-up (HR 0.98 per 10-year increase). Thus, the similar diagnostic yield of HCM in relatives across probands ages at diagnosis justifies family screening regardless of the proband age at diagnosis.

### Baseline characteristics in probands according to age at diagnosis

At baseline, we observed that HCM probands diagnosed >65 years had less LVH, higher LVOT gradients and more hypertension compared to those diagnosed at younger ages. Previous studies have shown that hypertension and obesity were associated with higher provocable LVOT gradients and LVH ^19^, suggesting that hypertension and obesity could contribute to the presentation of the HCM phenotype and LVOT obstruction in older probands. In our study, we found that HCM probands diagnosed at >65 years also had significantly more dyspnea and larger indexed left atrial volume at baseline compared to probands diagnosed at younger ages. Consistent with findings from Ljungman et al ^20^ who reported that HCM patients later in life often present with a greater symptom burden and larger left atrial volumes, our results also suggest that older HCM patients may have comorbidities that require distinct management strategies.

### Genetic findings in probands according to age at diagnosis

A lower proportion of probands diagnosed at older ages carried LP/P genetic variants compared to probands diagnosed at younger ages. Our findings corroborate prior research showing that probands with sarcomere-negative HCM present later in life compared to patients with sarcomere mutations. ^21, 22^ Canepa et al reported that rapidly expanding international HCM populations include increasingly older patients with more frequent genotype-negative status ^6^, suggesting monogenic causes of HCM being more common in HCM patients diagnosed younger. This is also supported by a recent study indicating that isolated low-penetrance sarcomere variants are linked to an older age at HCM diagnosis and contribute to age-related remodeling, even in the absence of overt HCM. ^23^ Thus, comorbidities and a polygenic burden may, to a larger extent, contribute to HCM in older probands. ^24, 25^

### Clinical events in probands according to age at diagnosis

We observed distinct risk profiles in probands according to age at diagnosis. Probands diagnosed at an older age had a higher prevalence of LV obstruction, atrial fibrillation and all-cause mortality, while probands diagnosed younger had a higher prevalence of ventricular arrhythmias, ICD implantation and heart transplantation. Consistent with findings from Maron et al. who found that HCM patients diagnosed ≥60 years of age had a low risk for disease-related mortality ^10^, we found that, despite a higher all-cause mortality in probands diagnosed older, cardiovascular mortality was similar among probands diagnosed at different ages.

### Diagnostic yield of family screening

The diagnostic yield of family screening was higher at baseline screening in relatives of probands diagnosed younger and in genotype-positive families compared to gene-elusive cases, consistent with prior research. ^27^ The prevalence of HCM at baseline, when adjusting for relatives’ age at baseline, was higher in relatives of probands diagnosed younger up to relatives aged 45-65 years. Conversely, no difference in the diagnostic yield during follow-up screening or the incidence of clinical adverse events were observed according to the proband’s age at diagnosis. These results were consistent across both gene-positive and in gene-elusive families, aligning with previous studies that found similar HCM penetrance across sarcomere variant carriers regardless of age. ^28^ The results of our study underscore the importance of genetic counselling and family screening strategies in families of HCM probands regardless of age at diagnosis. This is notable, considering current guidelines on screening of relatives and follow-up of patients do not take the proband’s age at diagnosis into account. ^12^

### Study limitations

The study cohort was drawn from a regional assembly of tertiary centers with possible referral bias favoring younger probands or families with severe disease or strong HCM history, possibly having led to an overestimation of clinical yield, but findings likely reflect real-world settings with a high family screening rate at 90%. The retrospective design introduces potential unknown confounders and ascertainment bias, limiting generalizability. Additionally, the study’s focus on Danish citizens may also restrict generalizability to other populations with differing genetic and environmental factors influencing HCM. Moreover, this study did not adjust for family history of HCM, limiting the ability to evaluate the diagnostic yield in gene-elusive families consisting of several HCM cases compared to gene-elusive families consisting of few HCM cases.

### Conclusions

This study highlights a significant impact of proband age at diagnosis on clinical presentation, genetic findings, and outcomes. Probands diagnosed at older age exhibited a lower yield of genetic testing, and less ventricular hypertrophy, but were more symptomatic. Despite an association between probands diagnosed younger and higher diagnostic yield in family screening, the combined prevalence of HCM at baseline and the incidence of new cases during follow-up were similar among probands diagnosed at different ages, supporting family screening regardless of the age of the proband at diagnosis. Our findings suggest that HCM diagnosed at older ages may be influenced by comorbidities or a high polygenic burden rather than monogenic mutations. Future research should further explore the genetic and environmental contributions to late-onset HCM and refine risk stratification strategies for affected families.

### Funding support and author disclosures

The study was supported by Rigshospitalet Research Council. The funder had no part in the design of the study, the collection, analysis, or interpretation of data or publication. The authors have no disclosures to declare.

## Data Availability

Data is not publically available, but de-identified data will be available upon reasonable request.

